# Mapping the specificity of H3N2 strain-specific and cross-reactive human neutralizing antibodies elicited by the 2025-2026 influenza vaccine

**DOI:** 10.64898/2026.02.20.26346746

**Authors:** Jiaojiao Liu, Sydney Gang, Caroline Kikawa, Alesandra J. Rodriguez, Shuk Hang Li, Naiqing Ye, Tachianna Griffiths, Elizabeth M Drapeau, Reilly K. Atkinson, Andrea N. Loes, Ronald G. Collman, James A. Ferguson, Julianna Han, Andrew B. Ward, Jesse D. Bloom, Scott E. Hensley

## Abstract

An H3N2 variant, named subclade K, continues to circulate widely during the 2025-2026 influenza season. This virus possesses a hemagglutinin (HA) protein that has eleven substitutions relative to the HA of the Northern Hemisphere 2025-2026 H3N2 vaccine strain. Many of these substitutions are in epitopes in well-characterized HA antigenic sites. Despite this, interim vaccine effectiveness studies indicate that the 2025-2026 influenza vaccine provides moderate protection against H3N2 subclade K infection. We previously reported that many individuals who received the 2025-2026 influenza vaccine produced antibodies that inhibit H3N2 subclade K virus cellular attachment. Here, we show these individuals also produced antibodies that neutralize H3N2 subclade K virus infection, and we observed a strong correlation between hemagglutination-inhibition titers and neutralizing antibody titers. We completed additional specificity studies using samples from individuals who did or did not have antibodies that cross-reacted to H3N2 subclade K viruses. Using high-throughput neutralization assays, we determined that antibodies that bound to the vaccine strain but not H3N2 subclade K viruses typically targeted antigenic site B of HA. Conversely, we found that cross-reactive neutralizing antibodies elicited by vaccination commonly targeted antigenic site A, D, and E of HA that are conserved between the vaccine strain and H3N2 subclade K viruses. Additional electron microscopy-based polyclonal epitope mapping studies confirmed that cross-reactive antibodies elicited by vaccination typically target epitopes on the side of HA. Together, our studies provide an immunological explanation of why the 2025-2026 influenza vaccine was partially effective against antigenically advance H3N2 subclade K viruses. Our data suggest that vaccine strains for subsequent seasons need to be carefully considered, since subclade K viruses have already started to acquire additional substitutions in HA antigenic sites targeted by cross-reactive antibodies.

## Introduction

It is important to continuously monitor seasonal influenza virus antigenic evolution, so that vaccine strains can be updated in subsequent influenza seasons. An antigenic variant of H3N2 (named subclade K) emerged in 2025 and continues to circulate widely in many areas of the world^1^. The hemagglutinin (HA) protein of this variant differs from the HA of the Northern Hemisphere 2025-2026 H3N2 vaccine strain (A/Croatia/10136RV/2023) in eleven residues (K2N, T135K, S144N, N145S, N158D, I160K, Q173R, A186D, K189R, T328A, HA2: S49N). Many of the HA substitutions are in important antigenic sites, and ferrets exposed to the 2025-2026 H3N2 vaccine strain produce antibodies that poorly react to subclade K viruses^2^. Nonetheless, several interim vaccine effectiveness studies suggest that the 2025-2026 influenza vaccine provides moderate protection against subclade K virus infection in humans^3–5^. Consistent with this, we previously completed hemagglutination inhibition (HAI) assays and found that the 2025-2026 influenza vaccine elicits antibodies in many individuals that can prevent viral attachment of subclade K viruses^6^. In that study, we found that only 11% (8 of 76) of participants in our study were seropositive (≥40 HAI titer) against the A/New York/GKISBBBG87773/2025 subclade K virus before vaccination, and that seropositivity increased to 39% (30 of 76) after vaccination.

In this report, we completed additional experiments using the same samples that we previously^6^ used for subclade K HAI testing. We completed HAI assays with additional variant viruses, and we also performed *in vitro* neutralization assays. We found that some individuals possessed serum antibodies that potently neutralized subclade K viruses, while other individuals possessed mostly strain-specific antibodies that did not recognize subclade K viruses. We completed additional high-throughput neutralization assays and electron microscopy-based polyclonal epitope (EMPEM) studies to map the specificity of strain-specific and cross-reactive H3N2 antibodies in a subset of sera samples.

## Methods

### Human serum samples

Serum and plasma samples were collected from 76 healthy adults before and 27-30 days after receiving a standard dose of the egg-based 2025-2026 Flulaval Trivalent influenza vaccine (GaxoSmithKline) between October-November 2025. This study was approved by the Institutional Review Board of the University of Pennsylvania.

### Viruses

Influenza viruses were created by reverse genetics system as previously described^7,8^. The hemagglutinin (HA) and neuraminidase (NA) sequences for influenza viruses A/Croatia/10136RV/2023 (2025-2026 H3N2 vaccine strain, subclade J.2, GISAID accession number EPI_ISL_19296516), A/District Of Columbia/27/2023 (subclade J.2, GISAID accession number EPI_ISL_18937823), A/Colombia/1851/2024 (subclade J.2.3, GISAID accession number EPI_ISL_19727802), A/Sydney/1359/2024 (subclade J.2.4, GISAID accession number EPI_ISL_19711425) and A/New York/GKISBBBG87773/2025 (subclade K, GISAID accession number EPI_ISL_20126669) were synthesized and cloned into the pHW2000 plasmid (Twist Bioscience). Viruses were launched by transient transfection of HA, NA plasmids and 6 reverse genetics pHW2000 plasmids that encode the internal segments of A/Puerto Rico/8/1934 in co-culture MDCK-SIAT1 and 293T cells. The 2025-2026 H3N2 vaccine strain was then propagated in 10-day-old fertilized chicken eggs, and other four influenza viruses were passaged twice in MDCK-SIAT1 cells. All virus stocks were sequence confirmed after propagation and stored at - 80 °C.

### Focus reduction neutralization test (FRNT)

FRNT assays were completed as previously described^7–9^. Human serum samples were treated with receptor destroying enzyme (RDE) (Denka-Seiken) for 2 hours at 37 °C and then inactivated for 30 minutes at 56 °C. One day before the assay, 100 μL of MDCK-SIAT1 cells were seeded into the flat-bottom 96-well plates at the concentration of 2.5 × 10^5^/mL with overnight culture at 37°C in 5% CO_2_. RDE-treated serum samples were 2-fold serially diluted in serum-free Minimum Essential Media (MEM, Gibco) in round-bottom 96-well plates and then incubated with the same volume of ∼ 300 focus-forming units (FFUs) of virus for 1 hour at room temperature. MDCK-SIAT1 cells that were previously seeded in 96-well plates were then washed twice with serum-free MEM. After 1 hour incubation, 100 μL of serum-virus mixtures were added to each well of 96-well plates with confluent monolayers of washed MDCK-SIAT1 cells. After another hour incubation at 37°C in 5% CO_2_, cells were washed twice with serum-free MEM and then we added 1.25% Avicel in MEM supplemented with 50 μg/mL gentamicin-sulfate (Gibco) and 5 mM HEPES buffer (Gibco). Plates were incubated for 18 hours at 37°C in 5% CO_2_. Media was then removed and cells were fixed with 4% paraformaldehyde for 1 hour at 4°C. Cells were then permeabilized with 0.5% Triton X-100 for 7 minutes, and then blocked with 5% milk in PBS for 1 hour at room temperature. Plates were washed with distilled water 5 times after blocking, and a mouse monoclonal anti-influenza A virus nucleoprotein antibody (clone IC5-1B7) and then a peroxidase-conjugated goat anti-mouse antibody (Southern Biotechnology) were each diluted in blocking buffer and incubated on plates for 1 hour at room temperature after each other. Plates were washed 5 times after incubation with primary and secondary antibodies. For visualizing infected cells, TrueBlue TMB substrate (KPL) was added to plates and incubated for 1 hour in the dark for foci development. Plates were imaged and foci were quantified by an ELISpot reader. FRNT_90_ titers were recorded as the inverse of the highest dilution of sera that reduced the number of foci by at least 90%, based on the “virus only” control wells on each plate. We completed at least two independent replicates with each serum sample.

### Hemagglutinin inhibition assay (HAI)

HAI were performed as previously described^10^. Human serum samples were treated with RDE buffer as described above in the FRNT section. RDE-treated serum was then incubated with a 1.1 volume 10% (v/v) fresh turkey red blood cell (RBC) solution for 1 hour at 4 °C. RDE- and RBC-treated serum samples were then transferred and collected after centrifugation at 1200rpm for 10 minutes. Sera were 2-fold serially diluted in round-bottom 96-well plates with a 1:20 starting dilution, and four agglutinating doses of viruses were added in a total volume of 100 μL. Then 12.5 μL of a 2% (v/v) turkey RBC solution was added to each well. Agglutination was read out after 1 hour incubation at room temperature. HAI titers were defined as the reciprocal of the highest dilution that inhibited the hemagglutination of turkey red blood cells. For sera with a titer of less than 1:20, we assigned an HAI titer of 1:10. Two independent replicates were completed with each serum sample.

### Sequencing-based neutralization assays

We measured neutralization titers to a set of influenza viruses with HA proteins from recent and current human seasonal H3N2 strains using sequencing-based neutralization assays as previously described.^11^ The titers analyzed in depth here are a subset of those reported in that larger survey study. See Kikawa et al^11^ for full experimental details; the raw titers and data analysis pipeline are publicly available at https://github.com/jbloomlab/flu-seqneut-2025to2026. Briefly, these assays involve creating a library of barcoded influenza virions encoding HA ectodomains from the recent seasonal strains followed by identifying 16-nucleotide barcodes, with the other genes from the lab-adapted A/WSN/1933 strain. The viral library is incubated with serum dilutions, and then infected into cells with the infectivity of different viral variants quantified by sequence of the barcodes. See Loes et al^12^ and a step-by-step protocol on protocols.io (dx.doi.org/10.17504/protocols.io.kqdg3xdmpg25/v2) for additional details.

### EMPEM studies

EMPEM protocols were performed as described previously^13^. Plasma were heat inactivated in a 56°C water bath for 60 min and incubated on CaptureSelect IgG-Fc resin (Thermo Scientific) in a 1:1 volume ratio of serum to resin overnight. IgG-depleted sera were removed, and the resin was washed three times with 5 column volumes(CV) of 1 × PBS. IgG was eluted by incubation with 10 CV 0.1M glycine pH 2.0 buffer for 20 min followed by neutralization with 1M Tris-HCl pH 8 buffer, repeated twice. Samples were buffer exchanged into 1 × PBS using centrifugation with Amicon concentrators. Total IgG was digested with activated papain in digestion buffer (20 mM sodium phosphate, 10 mM EDTA, 20 mM cysteine, pH 7.4) at 37°C for 5 h and was quenched with Iodacetamide. Samples were buffer exchanged into 1 × PBS and capture select was used to remove cleaved Fc domains. To prepare antigen-polyclonal Fab (pFab) complexes, our updated spin concentrator preparation method was used^14^. Briefly, 2ug of HA was mixed with 60ug of pFab and incubated overnight at room temperature. The unbound pFab was removed by washing with TBS in a 100kDa Amicon Ultra 0.5ml centrifugal filter then diluted to 20 μg/ml for staining with with 2% uranyl formate on negative stain electron microscope (nsEM) grids. Polyclonal Samples were imaged on at Talos F200c operating at −200 kV and ×73,000 magnification using a Ceta 16M detector at 2 Å/pixel. Images were processed using RELION-4^15^ and final analyses performed in Chimera^16^.

## Results

### The 2025-2026 influenza vaccine elicits antibodies that neutralize diverse H3N2 viruses

We collected sera from 76 individuals before and 27-30 days after receiving the egg-based 2025-2026 Flulaval Trivalent influenza vaccine. Our study population included participants with diverse birth years (birth year range: 1944-2001, median birth year 1971). We previously completed HAI assays with these sera and the 2025-2026 H3N2 vaccine strain (A/Croatia/10136RV/2023) and a H3N2 subclade K virus (A/New York/GKISBBBG87773/2025)^6^. We found that some participants produced antibodies that efficiently inhibited H3N2 subclade K virus, while other participants did not produce these types of antibodies after vaccination.

The A/Croatia/10136RV/2023 H3N2 vaccine strain is an egg-adapted subclade J.2 virus. To better characterize the antibody responses elicited by this vaccine strain, we completed HAI assays with additional H3N2 viruses, including A/District of Columbia/27/2023 (a J.2 cell-based vaccine strain), A/Colombia/1851/2024 (a J.2.3 virus), and A/Sydney/1359/2024 (a J.2.4 virus). Relative to the egg-adapted J.2 vaccine strain, the HA of the cell-based J.2 virus possesses an antigenic site B substitution (A186D) substitution, the HA of the J.2.3 possesses antigenic site A (N145S), antigenic site B (N158K, A186D, K189R) and HA2 (S49N) substitutions, and the J.2.4 virus possesses antigenic site A (T135K, N145S) and antigenic site B (A186D, K189R) substitutions (**Figure 1A**). We compared HAI titers using subclade J.2, J.2.3, and J.2.4 viruses with HAI titers we previously obtained^6^ using a subclade K virus that possesses substitutions in antigenic site A (T135K, S144N, N145S), antigenic site B (N158D, I160K, A186D, K189R), and antigenic site C (Q173R) (**Figure 1A**). Before vaccination HAI titers were significantly higher against the egg-based J.2 egg-adapted vaccine strain compared to the J.2 cell-based virus, J.2.3, J.2.4, and K viruses (**Figure 1B**). After vaccination, HAI titers against each virus increased significantly, but antibody levels remained higher against the vaccine strain relative to the other viral strains tested (**Figure 1B**).

**Figure 1:**
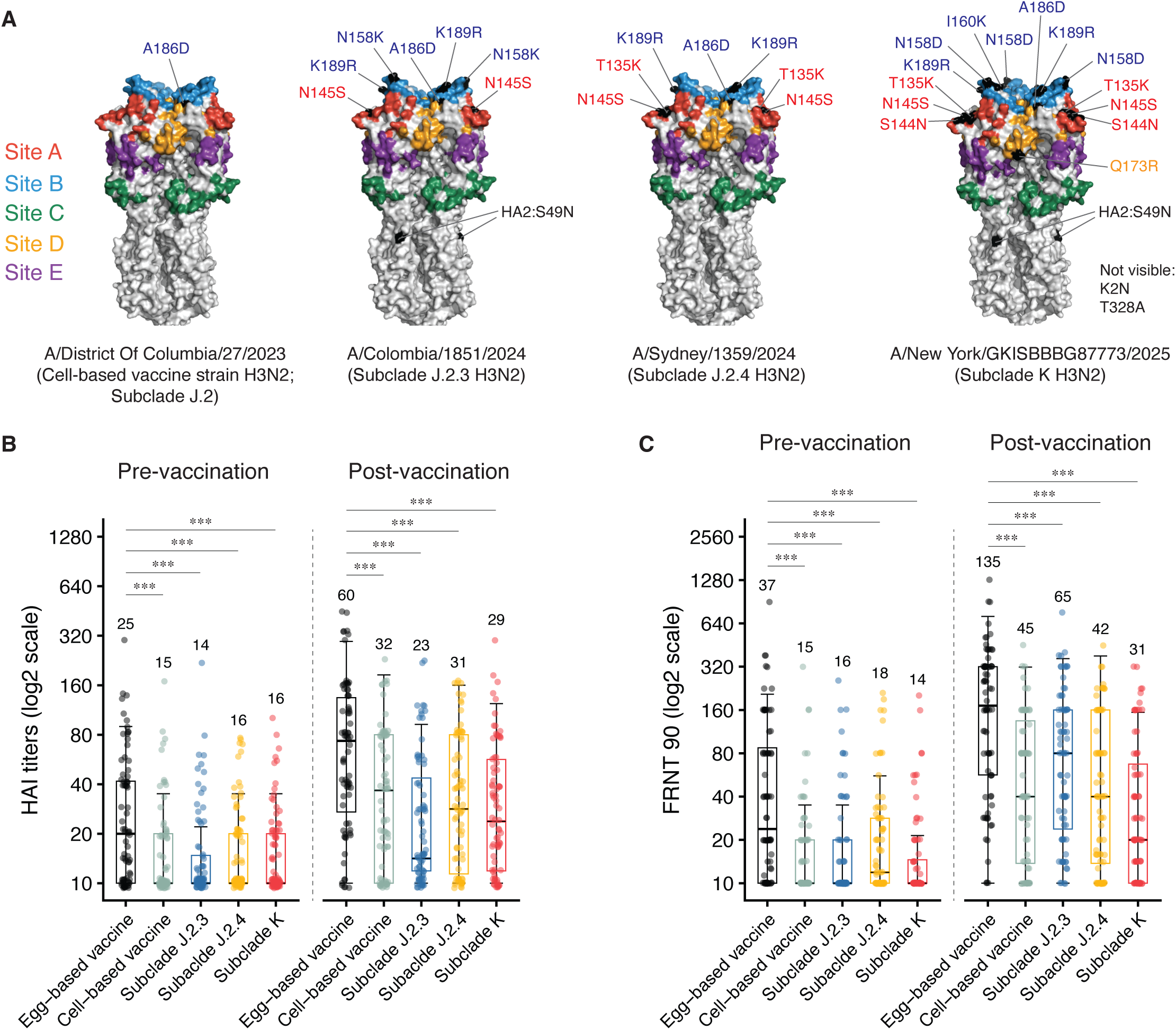
The 2025-2026 influenza virus vaccine elicits antibodies that neutralize diverse H3N2 viruses. (**A**) The crystal structure of the A/Victoria/22/2020 hemagglutinin (HA) trimer (PDB_8FAQ) with antigenic sites A-E are displayed. Amino acid differences between the HA of the 2025-2026 H3N2 egg-based vaccine strain (A/Croatia/10136RV/2023) and the 2025-2026 H3N2 cell-based vaccine strain (A/District Of Columbia/27/2023), H3N2 subclade J.2.3 virus (A/Colombia/1851/2024), H3N2 subclade J.2.4 virus (A/Sydney/1359/2024), and H3N2 subclade K virus (A/New York/GKISBBBG87773/2025) are shown in black. We completed hemagglutination inhibition (HAI) assays (**B**) and *in vitro* neutralization assays (**C**) using sera samples collected from 76 healthy adults before and after (27-30 days) after receiving a standard dose of the 2025-2026 egg-based influenza vaccine. HAI titers and *in vitro* neutralization titers (FRNT 90) against the 2025-2026 egg and cell-based H3N2 vaccine strains (subclade J.2), subclade J.2.3, subclade J.2.4 and subclade K H3N2 variant viruses are shown. Each dot in (**B**) and (**C**) represents the geometric mean titer using serum from one individual tested in two independent experiments. Boxes represent the interquartile range (IQR), with the center lines indicating the median titers. Whiskers extend to 1.5 × IQR. Geometric mean titers (GMT) is displayed above each box. The Wilcoxon Rank-Sum test was performed to compare HAI titers and neutralization titers (FRNT 90) between 2025-2026 egg-based H3N2 vaccine strain and each subclade variant strains. *** P < 0.001.

HAI assays measure antibodies that inhibit viral attachment. We completed additional *in vitro* neutralization assays since some antibodies inhibit viral replication through mechanisms independent of viral attachment. Consistent with our HAI data, neutralizing antibody titers were significantly higher against the J.2 egg-adapted vaccine strain compared to the other viral strains prior to vaccination (**Figure 1C**). Neutralizing antibody titers against each virus increased after vaccination, but like HAI titers, remained higher against the egg-adapted vaccine strain relative to the other viral strains tested (**Figure 1C**). HAI and *in vitro* neutralization titers correlated strongly (**Figure 2**), suggesting that most of the serum neutralizing antibodies block viral attachment.

**Figure 2:**
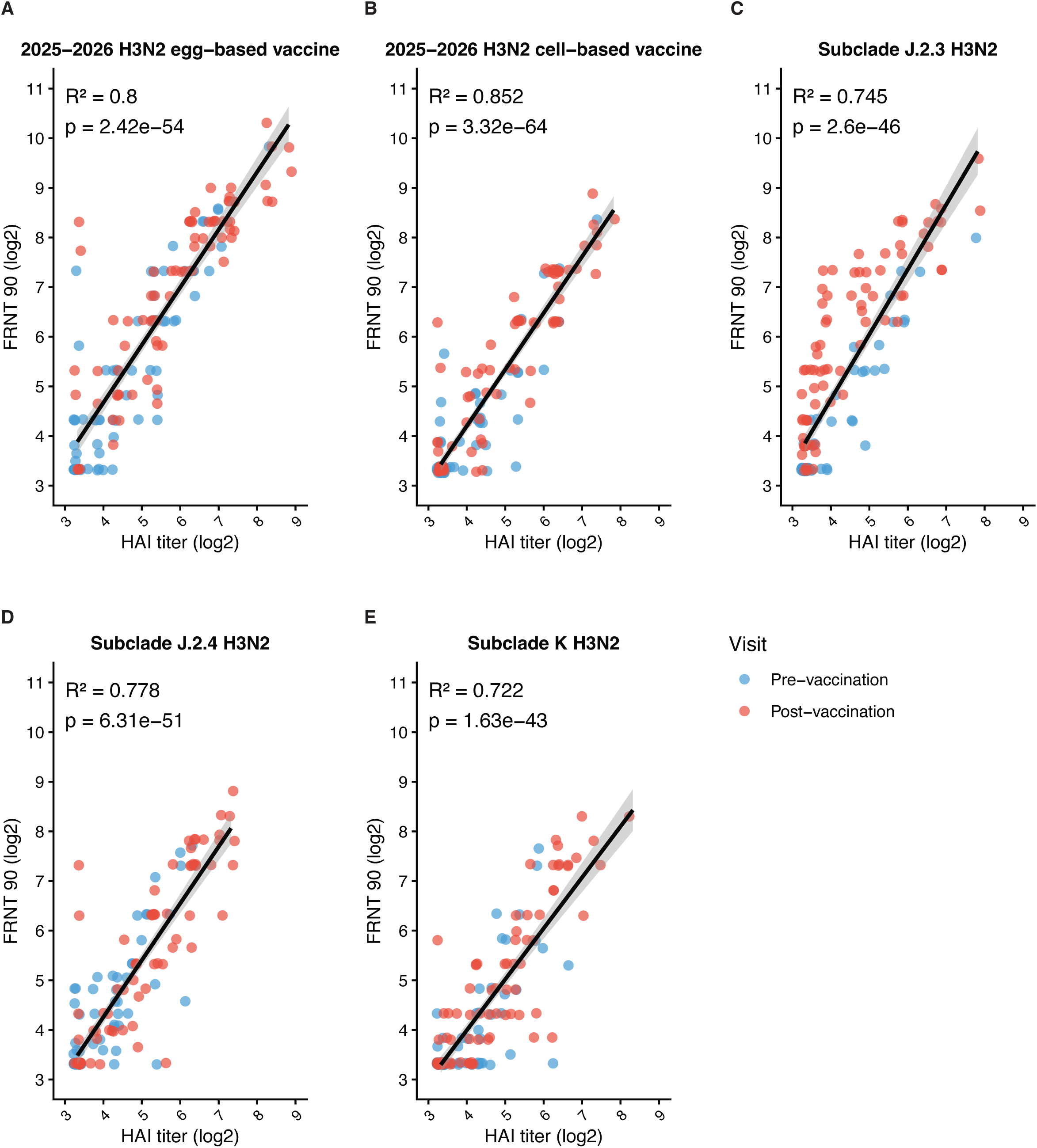
Most H3N2 neutralizing antibodies elicited by the 2025-2026 influenza vaccine block viral attachment. Panels **A** to **E** plot HAI titers versus neutralization titers (FRNT 90) of the data presented in Figure 1. Pre-vaccination samples are shown in blue and post-vaccination samples are shown in red. Each dot represents the log_2_-transformed geomean HAI titer and geomean neutralization titer of one individual from two independent experiments. The dark line represents the liner regression fit across all serum samples, 95% confidence intervals are shown as the shaded area. Pearson correlation coefficients (R^2^) and corresponding p-value were calculated by using liner regression.

### Specificity of H3 antibodies elicited by the 2025-2026 influenza vaccine

Since subclade K H3N2 viruses continue to circulate widely in humans, we completed additional studies to elucidate the specificity of antibodies against these viruses. We analyzed a subset of samples that had strain-specific antibodies that reacted to the J.2 vaccine virus but not the subclade K virus (herein, referred to as Category 1), and samples that had cross-reactive antibodies that recognized both J.2 and subclade K virus efficiently (herein, referred to as Category 2). For Category 1, we included 9 samples that had ≥4 fold reductions in HAI titers against subclade K viruses compared to titers against the egg-adapted J.2 vaccine strain (**Figure 3A**). For Category 2, we included 13 samples that had HAI titers of at least 80 against subclade K viruses (**Figure 3B**).

**Figure 3:**
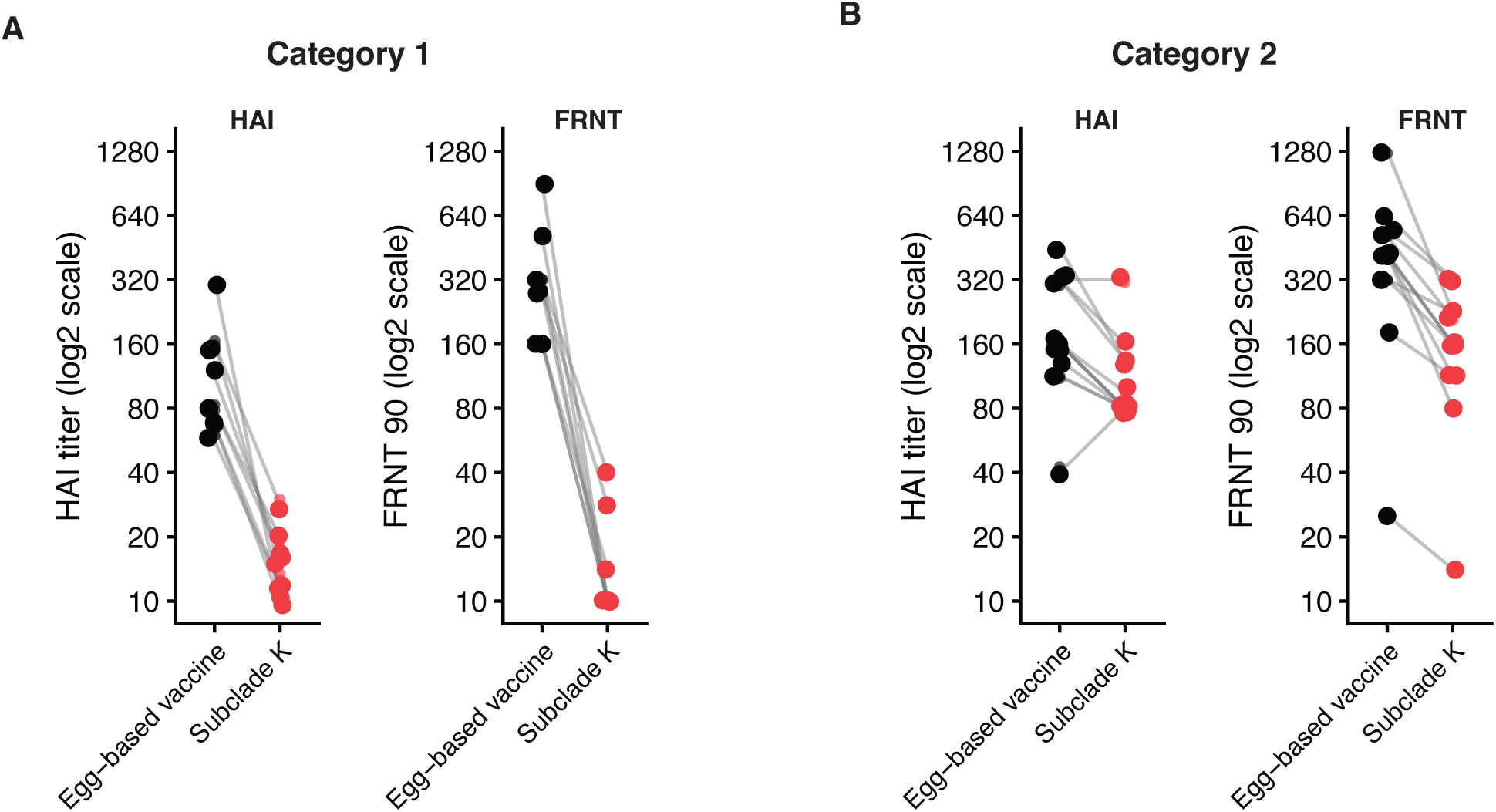
Selection of 2 groups of samples for further antigenic characterizations. We completed additional studies to define the specificity of antibodies in samples from a subset of individuals who had antibodies that strongly reacted to the J.2 vaccine virus but not subclade K (Category 1) and samples that had cross-reactive antibodies that recognized both J.2 and subclade K viruses (Category 2). Vaccine and subclade K HAI and *in vitro* neutralization titers of the Category 1 (**A**) and Category 2 (**B**) samples that we further characterized in Figures 4-6 are shown here.

We performed high-throughput sequencing-based neutralization assays using plasma from participants before and after vaccination. For these experiments, neutralization assays were completed using a pool of viruses that had unique barcodes, and we used sequencing fo the barcodes to reconstruct neutralization curves for each virus in the pool as previously described^12,17^. We included a library of HAs from 53 recently circulating human H3N2 strains and 30 H1N1 strains and the full data set from these experiments can be found in Kikawa et al.^12^. Here, we focused our more in-depth analyses on samples obtained from the 9 individuals in Category 1 and 13 individuals in Category 2. With each sample, we examined antibody reactivity to 10 subclade J.2 viruses (including the A/Croatia/10136RV/2023 egg-adapted virus), and 20 subclade K viruses.

Consistent with our conventional neutralization assays, samples from Category 1 participants neutralized the vaccine strain but failed to efficiently neutralize subclade K viruses (**Figure 4A**). These samples failed to efficiently recognize J.2 viruses with the A186D substitution in HA antigenic site B, suggesting that a large proportion of antibodies in these samples target the top of the variable HA head. It is also possible that the A186D substitution promoted viral escape by increasing affinity to cellular receptors, as previously described for H1N1 viruses^18^. Additional substitutions in antigenic sites A (N145S) and B (N158K, K189R) on the top of the HA head further reduced antibody recognition of samples from Category 1 participants. Conversely, samples from Category 2 participants efficiently neutralized both subclades J.2 and K viruses (**Figure 4B**). Serum antibodies from this group of participants were slightly affected by antigenic site A and B substitutions, but less so compared to samples from Category 1 participants. While antibodies from Category 2 participants effectively neutralized subclade K viruses, additional substitutions in HA antigenic site A (T135E), D (S96C and K207Q), and E (Q80K) reduced recognition of subclade K viruses (**Figure 4B**). This suggests that some participants have cross-reactive antibodies that target epitopes on the top and side of the HA of subclade K viruses.

**Figure 4:**
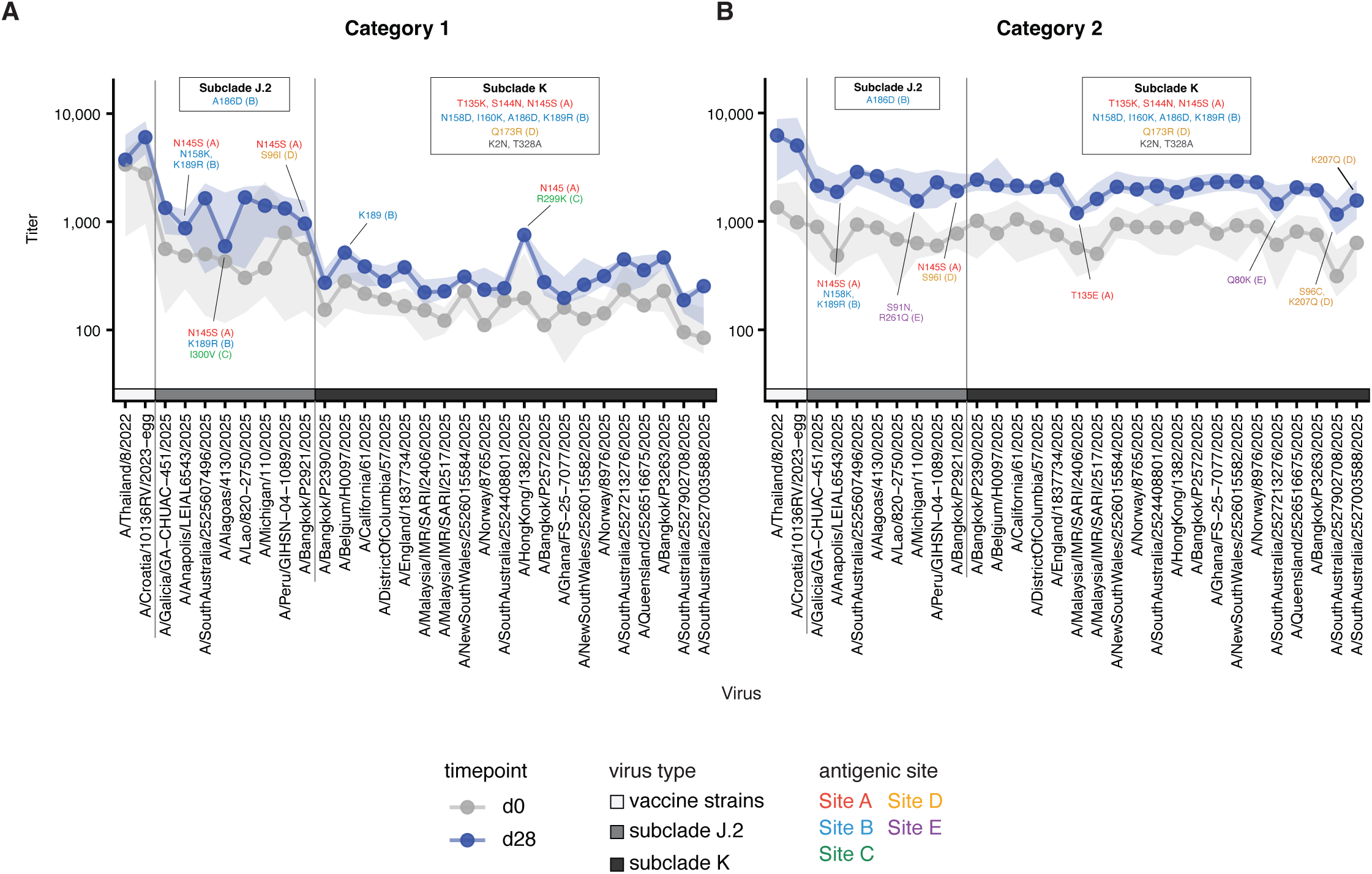
H3N2 vaccine strain-specific antibody responses typically target HA antigenic site B, whereas cross-reactive antibody responses target HA antigenic sites A, D, and E. We used high-throughput sequencing-based neutralization assays to measure antibody titers against 30 H3N2 viruses in plasma samples collected before (grey) and after (blue) vaccination of Category 1 (n=9) and Category 2 (n=13) participants. The grey and blue points show the median titers within the respective Category and timepoint, and the shaded area indicate the interquartile range. The 30 viruses are divided into three categories, denoted by the boxes along the x-axis and the vertical lines. The first two viral strains are the 2024-2025 Northern Hemisphere egg-based vaccine strain (A/Thailand/8/2022) and the 2025-2026 Northern Hemisphere egg-based vaccine strain (A/Croatia/10136RV/2023). The shared substitutions within the 8 subclade J.2 strains and 20 subclade K strains are shown above each subclade and each substitution is colored by antigenic site. For Category 1, the three J.2 strains with the lowest median titers and two K strains with the highest median titers within their respective subclades are annotated with substitutions present in antigenic site. For Category 2, the three J.2 and three K strains with the lowest median titers within their respective subclades are annotated.

Antibody responses in samples from Category 1 participants were fairly homogenous (**Supplemental Figure 1**); however, we observed a lot of heterogeneity in the specificity of cross-reactive antibodies in samples from Category 2 participants (**Supplemental Figure 2**). For example, Category 2 participants #2, 22, and 65 possessed HA antigenic site E antibodies that did not efficiently recognize a mutant virus with the Q80K HA substitution, whereas Category 2 participants #6, 7, 18, 44 possessed HA antigenic site A antibodies that did not efficiently recognize a mutant virus with the T135E HA substitution (an example of a site E-biased participant and a site A-biased participant are shown in **Figure 5**). Category 2 participants #2, 4, 6, 12, 18, 25, 44, and 65 produced HA antigenic site D antibodies with reduced reactivity to a mutant virus with the S96C and K207Q HA substitutions. Thus, there is substantial variation in the specificity of cross-reactive antibodies among different individuals who received the 2026-2026 seasonal influenza vaccine.

**Figure 5:**
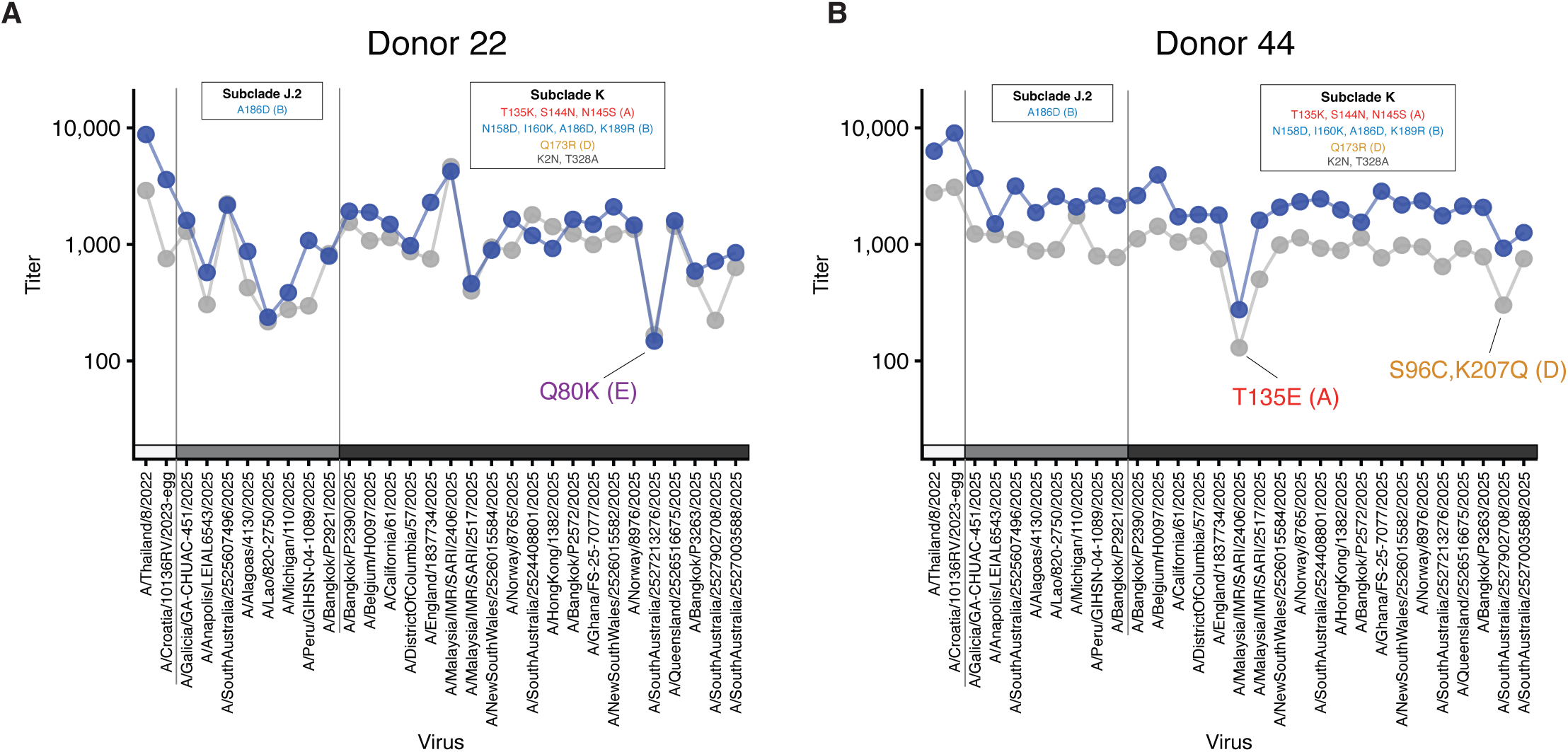
Different types of cross-reactive antibodies are enriched in different vaccinated participants. Pre-vaccination (grey) and post-vaccination (blue) antibody titers against the two most recent vaccine strains and a set of subclade J.2 and K strains are shown for Donors 22 and 44. The shared mutations within the J.2 strains and K strains are shown above each subclade and colored by antigenic site. Donor 22 has the lowest post-vaccination titer against A/SouthAustralia/2527213276/2025, which harbors a Q80K mutation in antigenic site E. Donor 44 has the lowest post-vaccination titers against A/Malaysia/IMR/SARI/2406/2025, which harbors a T135E mutation in antigenic site A, and A/SouthAustralia/2527902708/2025, which harbors S96C and K207Q mutations in antigenic site D.

Finally, we completed EMPEM studies to visualize HA binding interactions of plasma antibodies from a subset of Category 1 and 2 participants. We observed antibodies that bound to the HA stalk of both the H3 vaccine strain and subclade K virus in most samples (**Figure 6A**). We detected antibodies that reacted to top of the HA head of the H3 vaccine strain in 3 of the 7 Category 1 samples and 1 of the 4 Category 2 samples (**Figure 6A**). These antibodies did not bind to the subclade K virus. We detected antibodies that reacted to the side of the HA head of both the vaccine strain and subclade K strain in 2 of the 7 Category 1 samples (**Figure 6A**; note reduced occupancy observed to subclade K HA). Three of the 4 Category 2 samples had antibodies that bound to the side of the HA head of the subclade K virus (**Figure 6A**). We reconstructed 3D maps of plasma antibodies binding to 2 major antigenic sites (**Figure 6B-C**) that corroborate the findings from our high-throughput neutralization assays. We identified sera antibodies that bind to the HA of the vaccine strain at HA residues within antigenic site A and B (**Figure 6B**). These antibodies did not bind to the subclade K HA in these EMPEM experiments. We also identified sera antibodies from Category 2 participants that target epitopes on the side of HA that are conserved between the H3 vaccine strain and subclade K virus (**Figure 6C**).

**Figure 6:**
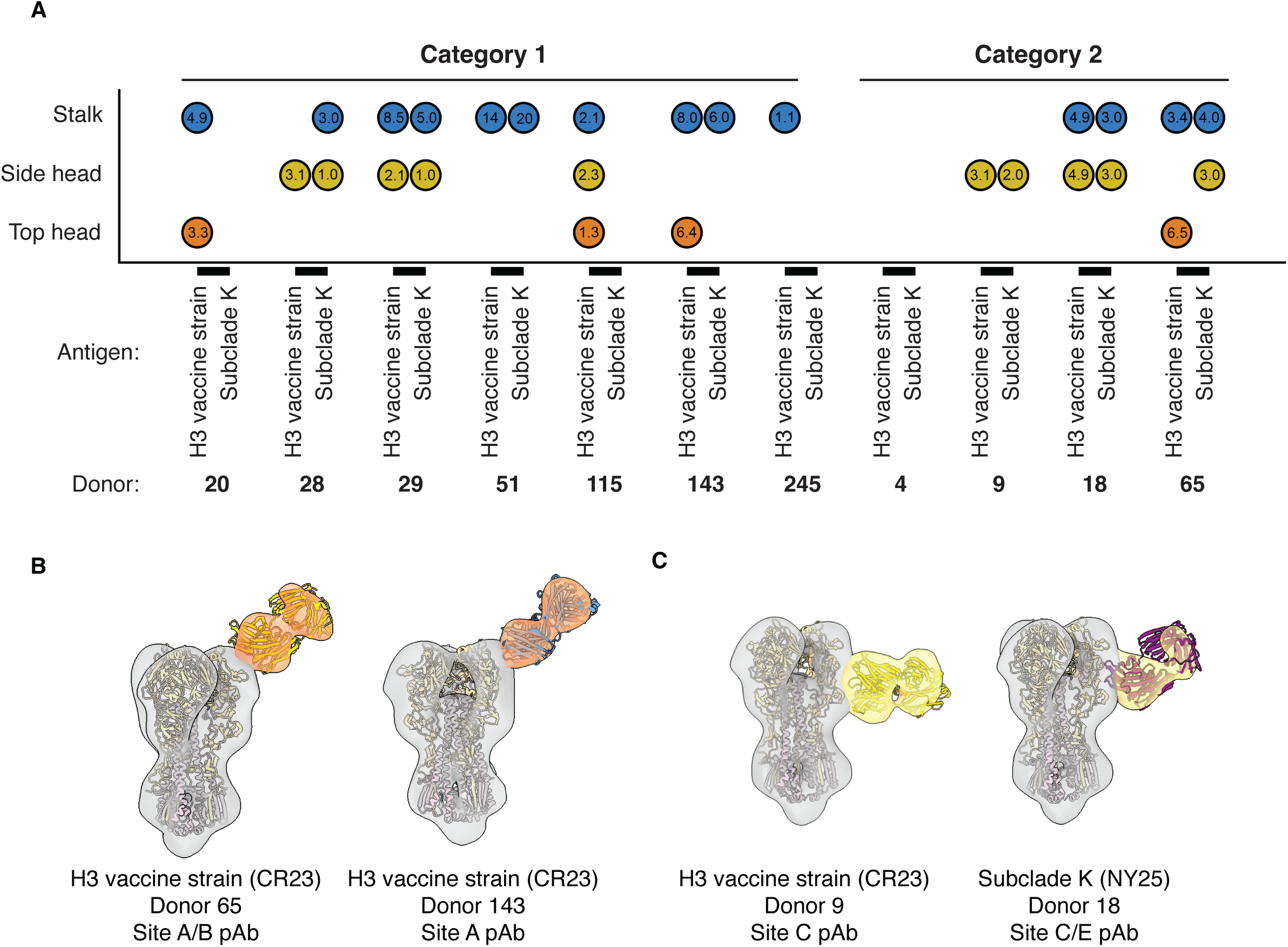
EMPEM epitope mapping of polyclonal antibodies that bind to the H3N2 vaccine strain and subclade K virus HAs. (**A**) Summary of EMPEM data using plasma samples from seven Category 1 and four Category 2 participants. Values inside circles are percentage of HA particles detected with antibodies to each HA epitope (stalk, side head, top head). (**B-C**) Representative negative stain EM reconstructions of polyclonal fab fragments binding to the top (**B**) or side (**C**) of the HA head from the A/Croatia/10136RV/2023 H3 vaccine strain and the A/New York/GKISBBBG87773/2025 subclade K virus.

These antibodies predominantly bind to residues within antigenic site C and E of HA. Ongoing studies are being completed to identify the binding footprints of additional sera samples that have different antibody specificities based on our high-throughput neutralization assays. For example, it will be important for us to further map the footprints of cross-reactive antibodies that bind to antigenic site A and antigenic D that were identified in our high-throughput neutralization assays.

## Conclusions

Our studies demonstrate that the egg-adapted 2025-2026 influenza vaccine that contains a subclade J.2 H3N2 antigen elicits neutralizing antibodies that cross-react to subclade J2.3, subclade J2.4, and subclade K HAs. We found that antibody responses varied greatly between different vaccinated participants in our study, with some individuals producing predominately antibodies that were vaccine strain-specific, while other individuals produced cross-reactive antibodies. We further show that there is great heterogeneity in the types of cross-reactive antibodies that different participants produced following vaccination. For example, some participants produced cross-reactive antibodies that predominantly targeted HA antigenic site A of subclade K viruses, whereas other participants produced cross-reactive antibodies that targeted HA antigenic site E of subclade K viruses (**Figure 5**).

The heterogeneity that we find in cross-reactive antibodies in this study is important, both from a basic science and translational perspective. From a basic science perspective, it will be interesting to determine why different humans mount antibodies of different specificities. Early childhood influenza exposures and immune history greatly influence the specificity of antibodies elicited by contemporary viral strains^19,20^, and it will be important for future studies to determine if cross-reactive HA site A versus site E antibodies are prevalent in individuals who have distinct birth years. From a translational perspective, the observed antibody heterogeneity in our study should be considered when new influenza vaccines are selected for the upcoming 2026-2027 season. It will be important that future vaccines are matched to circulating strains in major antigenic sites on the top of HA (like antigenic sites A and B), as well as other antigenic sites on the side of HA (like antigenic sites C, D, and E) that are typically thought to be more subdominant. For example, subclade K viruses with the Q80K HA site E substitution have been detected at relatively high frequencies in some parts of the world, and it is unclear if future vaccine strains with an Q80 or K80 HA should be selected for the 2026-2027 influenza season. Antibody responses against antigenic sites, such as site E, that are usually subdominant could potentially dominate antibody responses elicited by new subclade K vaccine strains in some individuals.

Our studies highlight how basic science studies can help inform the process of selecting vaccine strains. A better understanding of factors that lead to heterogeneity of antibody responses across different individuals will ultimately lead to improved viral forecasting and more effective influenza vaccines.

## Data Availability

All data produced in the present work are contained in the manuscript.

## Acknowledgement

We gratefully acknowledge the Authors from the originating laboratories responsible for obtaining the specimens and the submitting laboratories for generating the genetic sequence and metadata and sharing via the GISAID Initiative, on which the viruses from this study are based (**Table S1**).

---

**Supplementary Figure 1.**
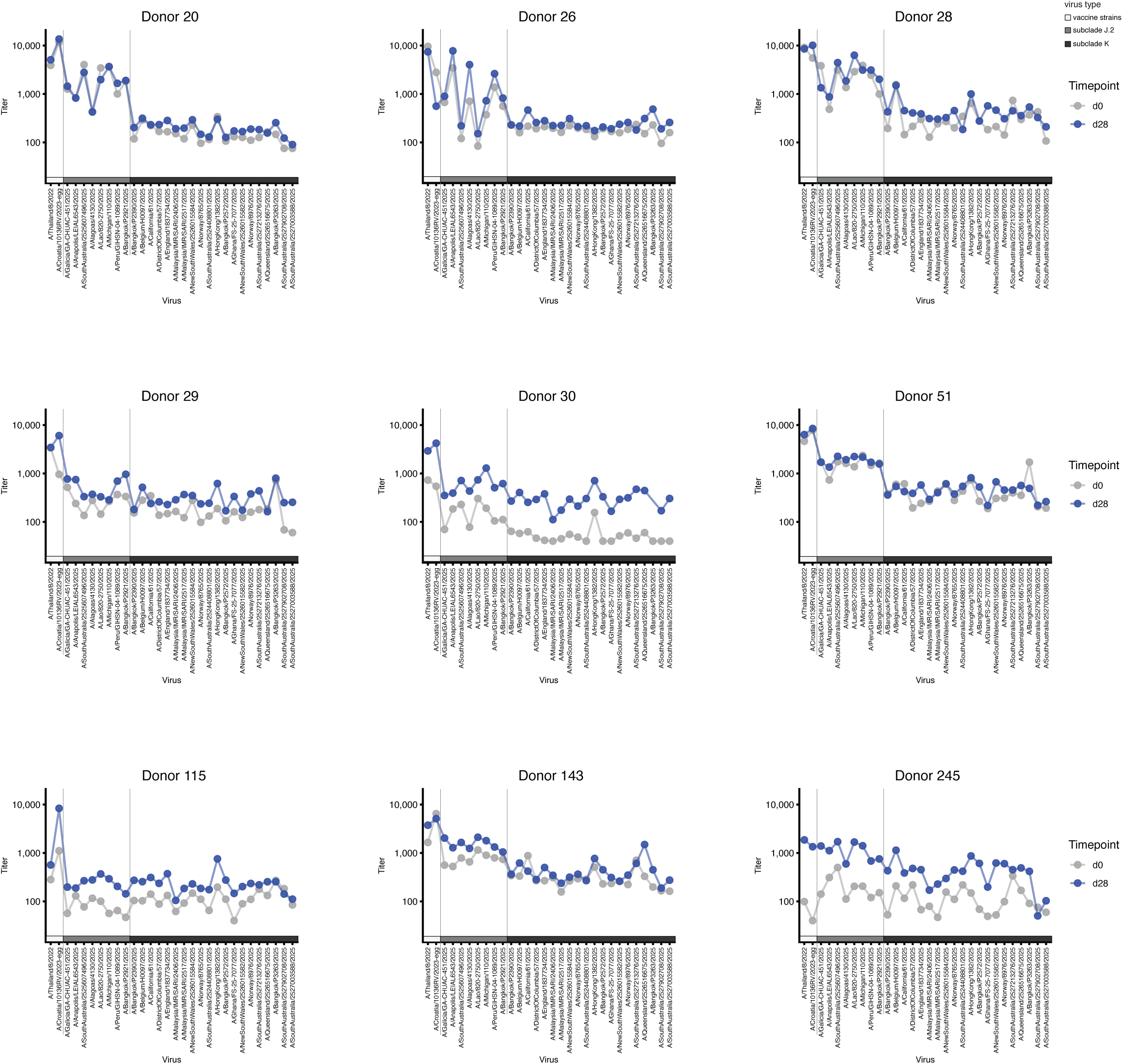

**Supplementary Figure 1.**
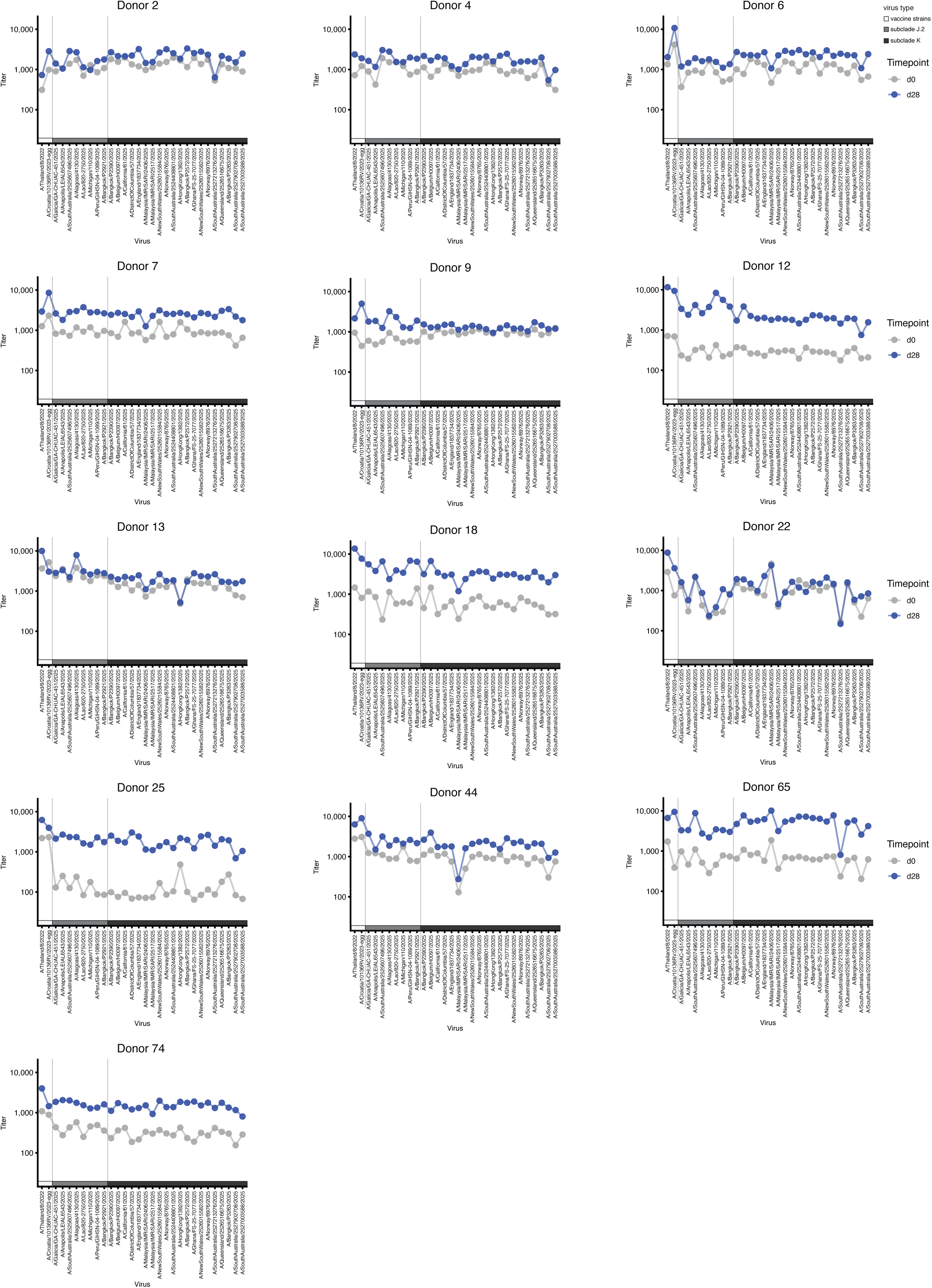

**Table S1:**
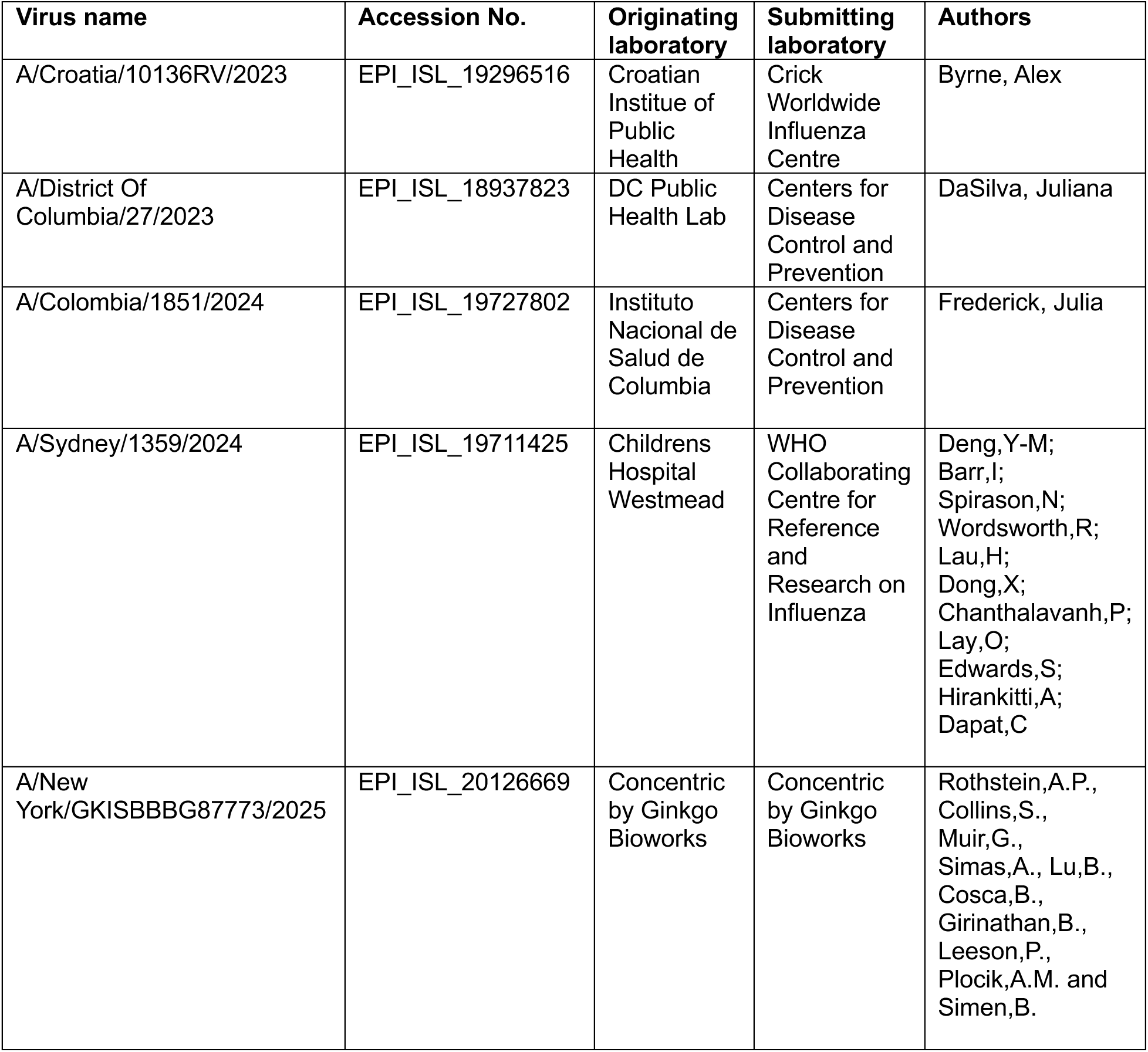
Acknowledgement to the contributors of the influenza virus sequences used in this study. We gratefully acknowledge the following Authors from the Originating laboratories responsible for obtaining the specimens and the Submitting laboratories for generating the genetic sequence and metadata and sharing via the GISAID Initiative, on which this research is based. All submitters of the data may be contacted directly via www.gisaid.org

